# Estimates of presumed population immunity to SARS-CoV-2 by state in the United States, August 2021

**DOI:** 10.1101/2021.09.17.21263759

**Authors:** Marie C.D. Stoner, Frederick J. Angulo, Sarah Rhea, Linda Morris Brown, Jessica E. Atwell, Jennifer L. Nguyen, John McLaughlin, David L. Swerdlow, Pia D.M. MacDonald

**Author notes:** Send all correspondence to: Marie Stoner. Women’s Global Health Imperative RTI International.

## Abstract

**Background:** Information is needed to monitor progress toward a level of population immunity to SARS-CoV-2 sufficient to disrupt viral transmission. We estimated the percentage of the United States (US) population with presumed immunity to SARS-CoV-2 due to vaccination, natural infection, or both as of August 26, 2021.

**Methods:** Publicly available data as of August 26, 2021, from the Centers for Disease Control and Prevention (CDC) were used to calculate presumed population immunity by state. Seroprevalence data were used to estimate the percentage of the population previously infected with SARS-CoV-2, with adjustments for underreporting. Vaccination coverage data for both fully and partially vaccinated persons were used to calculate presumed immunity from vaccination. Finally, we estimated the percentage of the total population in each state with presumed immunity to SARS-CoV-2, with a sensitivity analysis to account for waning immunity, and compared these estimates to a range of population immunity thresholds.

**Results:** Presumed population immunity varied among states (43.1% to 70.6%), with 19 states with 60% or less of their population having been infected or vaccinated. Four states have presumed immunity greater than thresholds estimated to be sufficient to disrupt transmission of less infectious variants (67%), and none were greater than the threshold estimated for more infectious variants (78% or higher).

**Conclusions:** The US remains a distance below the threshold sufficient to disrupt viral transmission, with some states remarkably low. As more infectious variants emerge, it is critical that vaccination efforts intensify across all states and ages for which the vaccines are approved.

**Summary:** As of August 26, 2021, no state has reached a population level of immunity thought to be sufficient to disrupt transmission. (78% or higher), with some states having remarkably low presumed immunity.

## INTRODUCTION

Closely monitoring the proportion of the population vaccinated against or previously infected with SARS-CoV-2 is critical for public health preparedness and response. As of August 22, 2021, 170.8 million people age 12 and older have been fully vaccinated and 37.6 million COVID-19 cases have been reported in the United States (U.S.).[1] Although the number of people who have been both vaccinated and had a reported COVID-19 infection is not known, vaccinated persons and those with reported infections represent 51.5% and 11.3%, respectively, of the total population.[1]

In mid-2021, the Delta variant (B.1.617.2), a World Health Organization variant of concern (VOC),[2] became the predominant SARS-CoV-2 variant circulating in the US. Associated with increased infectivity and severity,[2] Delta prevalence quickly increased from <1% of cases in May 2021 to >80% of cases in July 2021.[3] As more people are vaccinated or infected, immunity in the population will increase, approaching levels that may sufficiently slow community transmission. Population immunity, also referred to as herd immunity,[3] occurs when a sufficient proportion of a community is immune to a particular disease, making it unlikely for an infected person to come into contact with someone who is susceptible and thus substantially limiting pathogen transmission.[4,5] The level of immunity in a community that is sufficient to limit spread of a given pathogen is dependent on numerous factors, but is most simply calculated as 1-(1/R_0_), where R_0_ is the reproductive number or a measure of the degree to which the pathogen is contagious.[4,5] Population immunity threshold estimates for SARS-CoV-2 have ranged from 50% to 83%.[6–8] The threshold is likely even higher for variants that are more infectious than the original wild type, and therefore, have a higher reproductive number; Delta has an estimated population immunity threshold as high as 80%-90%.[9,10] Progress toward this threshold can be affected by the timing and extent of transmission patterns in different geographic areas. Thus, it is important to estimate how many people in a population have “presumed immunity” (i.e., have been infected or vaccinated) to SARS-CoV-2 to focus public health response efforts where they can achieve the biggest impact and thereby reduce risk of severe COVID-19 cases and associated deaths.

While several studies have speculated how close the US may be to reaching SARS-CoV-2 population immunity, few have attempted to robustly calculate presumed immunity percentages by state.[6,7,9,11,12] Population immunity to SARS-CoV-2 will result from both immunity from vaccination and immunity from natural infection; however, monitoring population immunity has largely been based on vaccination rates alone. Routine surveillance data on infections and vaccinations, while imperfect, can be used to provide an estimate of progress toward various immunity thresholds by estimating the percentages of vaccinated or infected persons. These data are dynamic, as is the required threshold, if VOCs with different R_0_ values continue to emerge. Therefore, it will be particularly important to continue to track this information as the pandemic progresses.

In this study, we estimated the percentage of the US population, by state, with presumed immunity to SARS-CoV-2 due to vaccination or natural infection (or a combination) as of August 26, 2021. Our study builds on other similar work through the use of seroprevalence surveys, which provide a more direct estimate of presumed immunity due to previous SARS-CoV-2 infection.[12] We used vaccination and seroprevalence survey data to derive estimates of the percentage of people who have presumed immunity due to either vaccination against COVID-19 or infection with SARS-CoV-2. To monitor progress toward population immunity, we then compared these estimates with a range of potential thresholds needed to achieve population-level immunity.

## METHODS AND MATERIALS

Using publicly available data, we employed a three-step process to calculate presumed population immunity by state (Figure 1). First, we calculated the percentage of the population who were presumed immune because of natural infection by using seroprevalence estimates by state and adjusting for underreporting. Second, we calculated the percentage of the population who were presumed immune because of vaccination by considering the vaccine effectiveness of being fully vaccinated (receiving the recommended doses for a COVID-19 vaccine) and being partially vaccinated (receiving only one dose of a two-dose vaccine). At the time of this analysis, federal guidelines did not include recommendation for booster vaccination(s) beyond the original dosing regimen specified by the vaccine manufacturers. Third, we estimated the percentage of the population for four mutually exclusive categories: (1) immune because of both vaccination and infection; (2) immune because of vaccination only; (3) immune because of infection only; and (4) not immune. Categories 1-3 were combined to create a group of individuals with “presumed immunity.” Our final estimate represents the total population by state meeting criteria for this composite category. Lastly, we compared these estimates with a range of potential values for the population immunity threshold to determine how close each state may be to achieving population immunity.[6–9,13]

**Figure 1.**
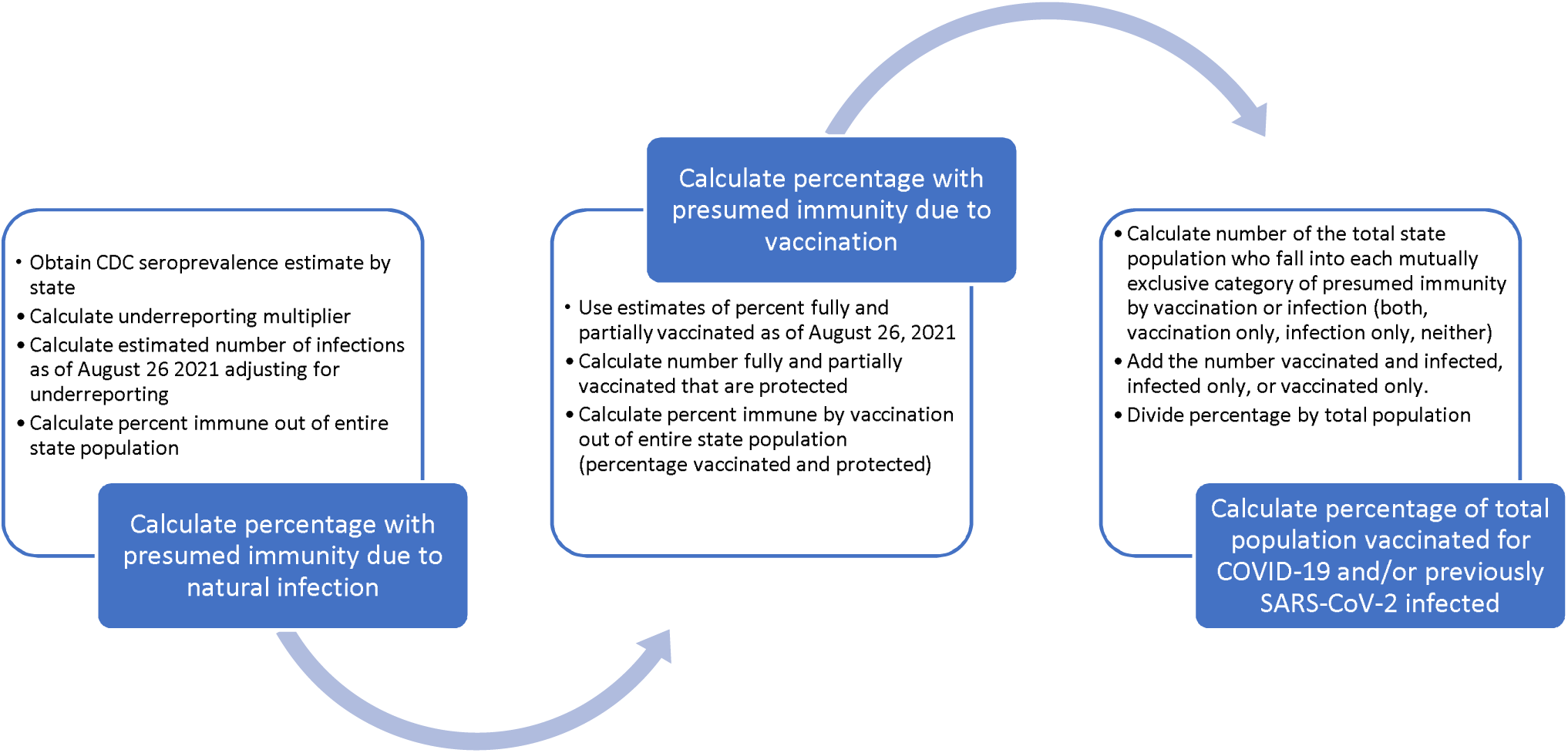
Process for calculating percentage of the population vaccinated for COVID-19 or previously SARS-CoV-2 infected by state as of August 26, 2021.

To estimate the number of people previously infected with SARS-CoV-2 by state, we followed Angulo and colleagues’ methods.[14] Specifically, we obtained the most recent seroprevalence data for a given state from the Centers for Disease Control and Prevention (CDC) Nationwide Commercial Laboratory Seroprevalence Surveys, which use blood samples (e.g., from routine medical care or sick visits) submitted to commercial laboratories for reasons unrelated to COVID-19 in all 50 U.S. states; Washington, DC; and Puerto Rico.[1,15,16] Of these blood samples, ∼1300/state are tested for SARS-CoV-2 every 2 weeks.[16] At the time of our analysis, the most recent estimates were from July 31, 2021. We included estimates from tests that used only the nucleocapsid protein as the viral antigen target rather than the spike protein because the nucleocapsid is a better representation of immunity due to infection.[1] To prevent unreliable estimates, we do not show results for states with seroprevalence estimates based on <1,000 samples. We multiplied the seroprevalence estimate for each state by the population of that state from the US Census[17] to derive an estimate of the number of persons infected on the date seroprevalence samples were collected. Next, we compared the estimated number of infected persons with the total number of cumulative reported infections on that date from CDC (June 31, 2021) to calculate the underreporting multiplier for that state. If the underreporting multiplier was <1, the multiplier was set at 1 to equal the number of reported cases. Lastly, we estimated the number of infections in the population on August 26, 2021, the most recent date at the time of analysis, by multiplying the cumulative number of infections on that day from CDC by the underreporting multiplier. We divided this number by the total population to derive the percentage of the total population with immunity due to infection as of August 26, 2021.

To determine how many people may have presumed immunity through vaccination, we used estimates of the percentage fully vaccinated and the percentage who had received at least one dose of a two-dose regimen, as reported by CDC on August 26, 2021.[1] We calculated the percentage immune from full vaccination by multiplying the total population fully vaccinated with each approved COVID-19 vaccine by the estimated protection against infection for Janssen (72% in US subsample[18,19]), Moderna (94%[20]), and Pfizer-BioNTech (95%[21]). We used an estimate of 85% protection for those vaccinated with an unknown COVID-19 vaccine.[22] We then calculated the number who had received only one dose of Moderna or Pfizer-BioNTech vaccine by subtracting those who were fully vaccinated from those with at least one dose and multiplied this number by estimated protection from one dose of a two-dose vaccine (82%)[23]. Next, we multiplied the percentages of persons immune from full vaccination (by any type of COVID-19 vaccine) and partial vaccination by the total population and added them together. We divided this number by the total population to estimate the percentage of the total population with vaccine-derived immunity as of August 26, 2021.

To determine the percentage of the total population in each state with some immunity due to vaccination, infection, or both, we defined immunity in the population based on four mutually exclusive categories: immune due to both vaccination and infection, immune due to vaccination only, immune due to infection only, and not immune. We calculated the percentage of the population in each category by multiplying the percentages with some immunity due to natural infection and vaccination. The four categories sum to 100% of the population. Finally, we added the percentages with immunity due to both infection and immunity, infection only, and vaccination only to get the percentage of the total population of each state with some immunity due to vaccination and/or infection as of August 26, 2021.

We conducted a sensitivity analysis to explore the effect of waning immunity from natural infection and from vaccination. For waning immunity from natural infection, we used CDC data[1] on the number of reported COVID-19 cases per day to estimate the proportion of the reported infections that occurred >12 months before August 26, 2021 (i.e., as of the end of August 2020), 6-12 months before August 26, 2021 (i.e., as of the end of February 2021), and <6 months before August 26, 2021.[1] For those infected during these time periods, we estimated immunity of 25%, 50%, and 100%, respectively. For waning immunity from vaccination, we used vaccine uptake data from the CDC[1] to estimate the proportion of the fully vaccinated who, as of August 26, 2021, had been fully vaccinated for ≥5 months (i.e., before the end of March 2021) and <5 months (i.e., after the end of March 2021).[1] For those who had been vaccinated for ≥5 months, we assumed vaccine efficacy of 65%.

The final estimate of the percentage of persons with presumed immunity due to COVID-19 vaccination or SARS-CoV-2 infection by state was compared with estimates of the population immunity threshold from the literature to assess each state’s progress toward population immunity. We used a range of potential thresholds, from a minimum of 67% to a maximum of 90% based on a range of R_0_ of the virus of 3 to 10, with the most likely number being ∼3 for early variants (67%) and 4.5 for more infectious variants (78%), although the R_0_ and corresponding threshold for the Delta variant is likely higher than 78%.[6–10,13,24,25] If future VOCs arise that are more transmissible, the R_0_ could be higher, leading to an even higher threshold.

## RESULTS

As of August 26, 2021, the percentage of the total population previously infected with SARS-CoV-2 by state ranged from 4.0% in Hawaii to 41.7% in Mississippi (Figure 2). The percentage vaccinated for COVID-19 ranged from 35.5% in Idaho to 61.5% in Massachusetts. Comparisons among the four categories indicated that the people with immunity due to vaccination only represented the largest proportion of the population (Table 1) and ranged from 22.1% in Mississippi to 57.7% in Hawaii. In the second category, the percentage of the population with presumed immunity due to both vaccination and infection ranged from 2.4% in Hawaii to 18.1% in Illinois and 18.0% in New Jersey. The states with the highest percentages of immunity due to both infection and vaccination were those where a large percentage of the population had been previously infected (e.g., 31.5% in New Jersey, 33.7% in Illinois). In the third category, the percentage with presumed immunity by infection only and not vaccination ranged from 1.6% in Hawaii to 25.9% in Mississippi. States with a high percentage of immunity due to infection only had relatively low or modest vaccination rates (e.g., 37.9% in Mississippi). In the fourth category, the percentage without immunity from either vaccination or infection ranged from 29.4% in New Jersey to 56.9% in Idaho. States with the highest percentages of the population without immunity had some of the lowest vaccination rates (e.g., 39.0% in West Virginia, 35.5% in Idaho,). In 19 states, ≥40% of the population had no immunity to SARS-CoV-2.

**Figure 2.**
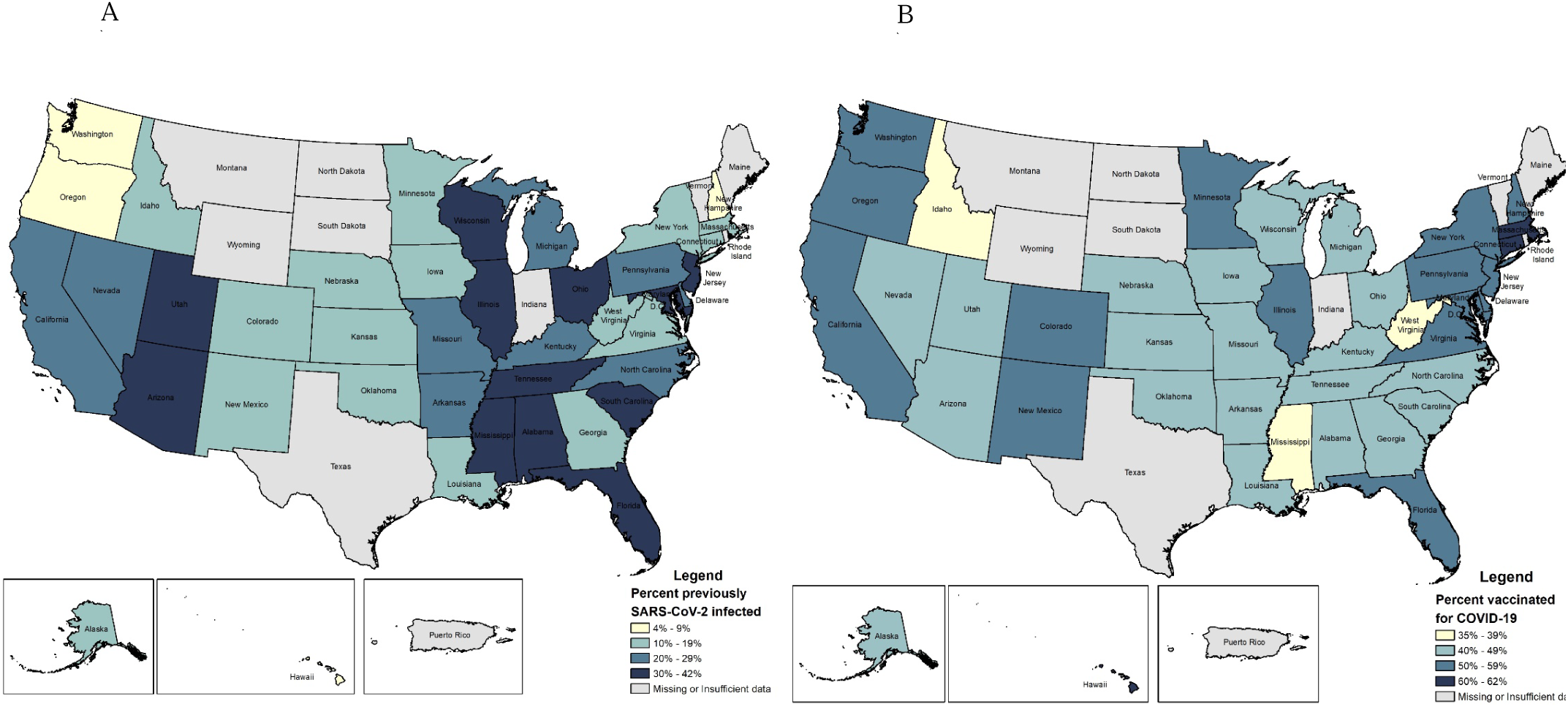
Percentage of the population (A) previously SARS-CoV-2 infected and (B) vaccinated for COVID-19 by state as of August 26, 2021.

**Table 1.** Percentage of the population in each category of presumed immunity due to natural infection, vaccination, both, or neither, by state as of August 26, 2021^a^.

| State<sup>b</sup> | Vaccination and<br>Infection, % | Vaccination and<br>No Infection, % | Infection and No<br>Vaccination, % | No Immunity,<br>% | Total Population<br>Vaccinated for COVID-19<br>and/or Previously SARS-CoV-2 Infected, % | State Rank |
| --- | --- | --- | --- | --- | --- | --- |
| Alabama | 13.6% | 26.8% | 20.0% | 39.6% | 60.4% | 23 |
| Alaska | 5.0% | 39.5% | 6.3% | 49.3% | 50.7% | 38 |
| Arizona | 14.8% | 31.5% | 17.1% | 36.5% | 63.5% | 12 |
| Arkansas | 12.6% | 31.1% | 16.3% | 40.0% | 60.0% | 24 |
| California | 12.7% | 43.5% | 9.9% | 33.9% | 66.1% | 6 |
| Colorado | 5.3% | 46.6% | 4.9% | 43.2% | 56.8% | 31 |
| Connecticut | 6.2% | 53.7% | 4.2% | 35.9% | 64.1% | 9 |
| Delaware | 11.9% | 40.5% | 10.8% | 36.7% | 63.3% | 13 |
| Florida | 15.9% | 35.9% | 14.8% | 33.4% | 66.6% | 5 |
| Georgia | 5.9% | 35.7% | 8.3% | 50.1% | 49.9% | 39 |
| Hawaii | 2.4% | 57.7% | 1.6% | 38.3% | 61.7% | 20 |
| Idaho | 4.2% | 31.3% | 7.6% | 56.9% | 43.1% | 42 |
| Illinois | 18.1% | 35.6% | 15.6% | 30.7% | 69.3% | 2 |
| Iowa | 6.2% | 40.2% | 7.2% | 46.5% | 53.5% | 34 |
| Kansas | 5.8% | 40.9% | 6.6% | 46.6% | 53.4% | 35 |
| Kentucky | 13.7% | 33.0% | 15.6% | 37.7% | 62.3% | 18 |
| Louisiana | 5.9% | 34.7% | 8.6% | 50.7% | 49.3% | 40 |
| Maryland | 17.0% | 38.5% | 13.7% | 30.8% | 69.2% | 3 |
| Massachusetts | 6.7% | 54.8% | 4.2% | 34.3% | 65.7% | 8 |
| Michigan | 13.2% | 32.1% | 15.9% | 38.8% | 61.2% | 21 |
| Minnesota | 5.9% | 44.6% | 5.8% | 43.6% | 56.4% | 32 |
| Mississippi | 15.8% | 22.1% | 25.9% | 36.2% | 63.8% | 10 |
| Missouri | 11.2% | 31.6% | 14.9% | 42.3% | 57.7% | 30 |
| Nebraska | 5.8% | 40.8% | 6.7% | 46.7% | 53.3% | 36 |
| Nevada | 13.6% | 33.7% | 15.2% | 37.6% | 62.4% | 17 |
| New Hampshire | 4.3% | 50.9% | 3.5% | 41.3% | 58.7% | 28 |
| New Jersey | 18.0% | 39.0% | 13.6% | 29.4% | 70.6% | 1 |
| New Mexico | 7.4% | 49.8% | 5.6% | 37.2% | 62.8% | 16 |
| New York | 7.1% | 47.9% | 5.8% | 39.2% | 60.8% | 22 |
| North Carolina | 11.4% | 33.8% | 13.8% | 41.1% | 58.9% | 27 |
| Ohio | 15.2% | 27.8% | 20.1% | 36.8% | 63.2% | 14 |
| Oklahoma | 6.0% | 37.6% | 7.7% | 48.7% | 51.3% | 37 |
| Oregon | 4.5% | 47.7% | 4.1% | 43.7% | 56.3% | 33 |
| Pennsylvania | 13.4% | 43.3% | 10.2% | 33.0% | 67.0% | 4 |
| South Carolina | 12.5% | 29.3% | 17.4% | 40.8% | 59.2% | 25 |
| Tennessee | 12.0% | 28.0% | 18.0% | 42.0% | 58.0% | 29 |
| Utah | 14.3% | 31.8% | 16.7% | 37.1% | 62.9% | 15 |
| Virginia | 9.9% | 43.1% | 8.8% | 38.1% | 61.9% | 19 |
| Washington | 4.5% | 50.9% | 3.6% | 41.1% | 58.9% | 26 |
| Washington, D.C. | 13.2% | 41.9% | 10.8% | 34.1% | 65.9% | 7 |
| West Virginia | 3.9% | 35.2% | 6.0% | 54.9% | 45.1% | 41 |
| Wisconsin | 14.4% | 33.6% | 15.6% | 36.4% | 63.6% | 11 |

<sup>a</sup>Puerto Rico and North Dakota are not included because of missing data on seroprevalence; Montana, Indiana, South Dakota, Maine, Rhode Island, Vermont, Texas, and Wyoming are not shown because of seroprevalence samples under 1000.
<sup>b</sup>, Estimates are for the total population, including all ages.

The overall percentage of each state with presumed immunity due to either vaccination or infection ranged from 43.1% in Idaho to 70.6% in New Jersey (Figure 3). Using a minimum threshold of 67% corresponding to an R_0_ of 3, only four states would have reached population immunity (New Jersey, Illinois, Maryland, Pennsylvania).[6–9,13] Based on a higher R_0_ of 4.5 for more infectious variants (78% threshold), no states would have reached the population immunity threshold as of August 26, 2021.[6–9,13] Given that the threshold for the Delta variant is likely even higher than 78%, we can assume that no states will have reached that threshold.[25]

**Figure 3.**
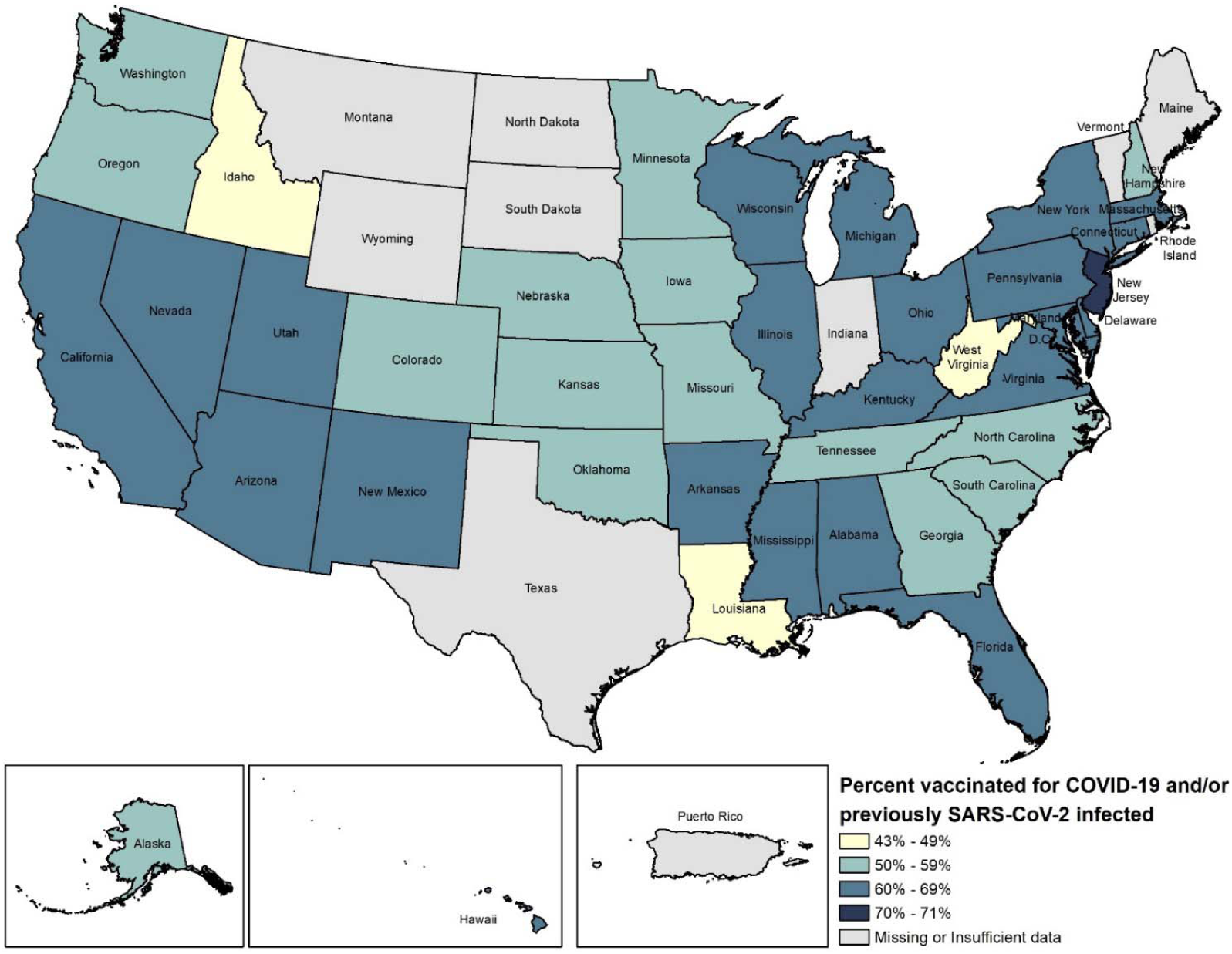
Percentage of the population vaccinated for COVID-19 and/or previously SARS-CoV-2 infected by state as of August 26, 2021.

In the sensitivity analysis assessing the impact of waning immunity, we estimated that the percentage with presumed immunity due to either vaccination or infection would be lower than originally calculated and would range from 37.9% in Idaho to 62.3% in New Jersey (Table 2), and none of the states would meet even the minimum threshold of 67%.

**Table 2.:**
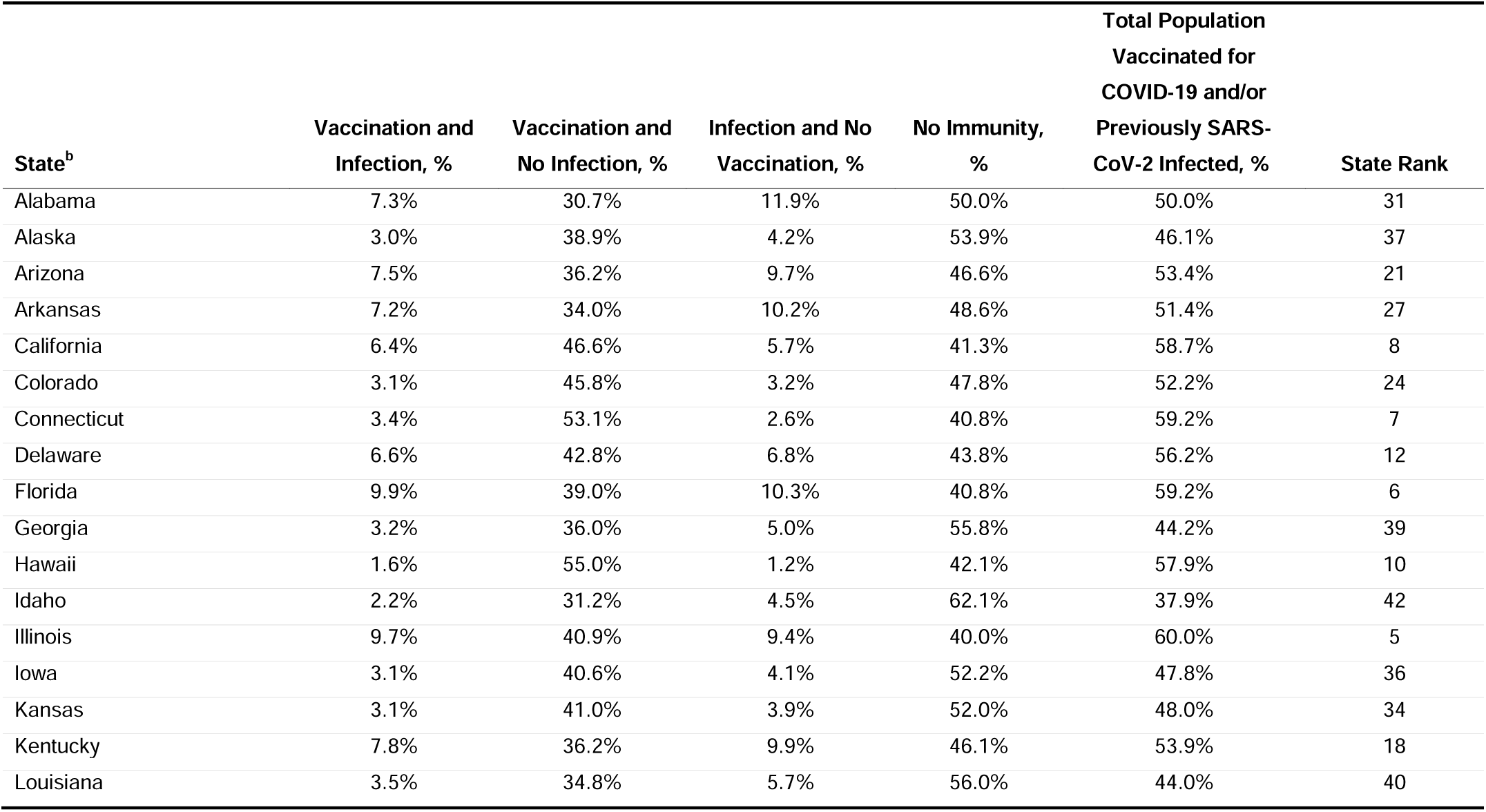
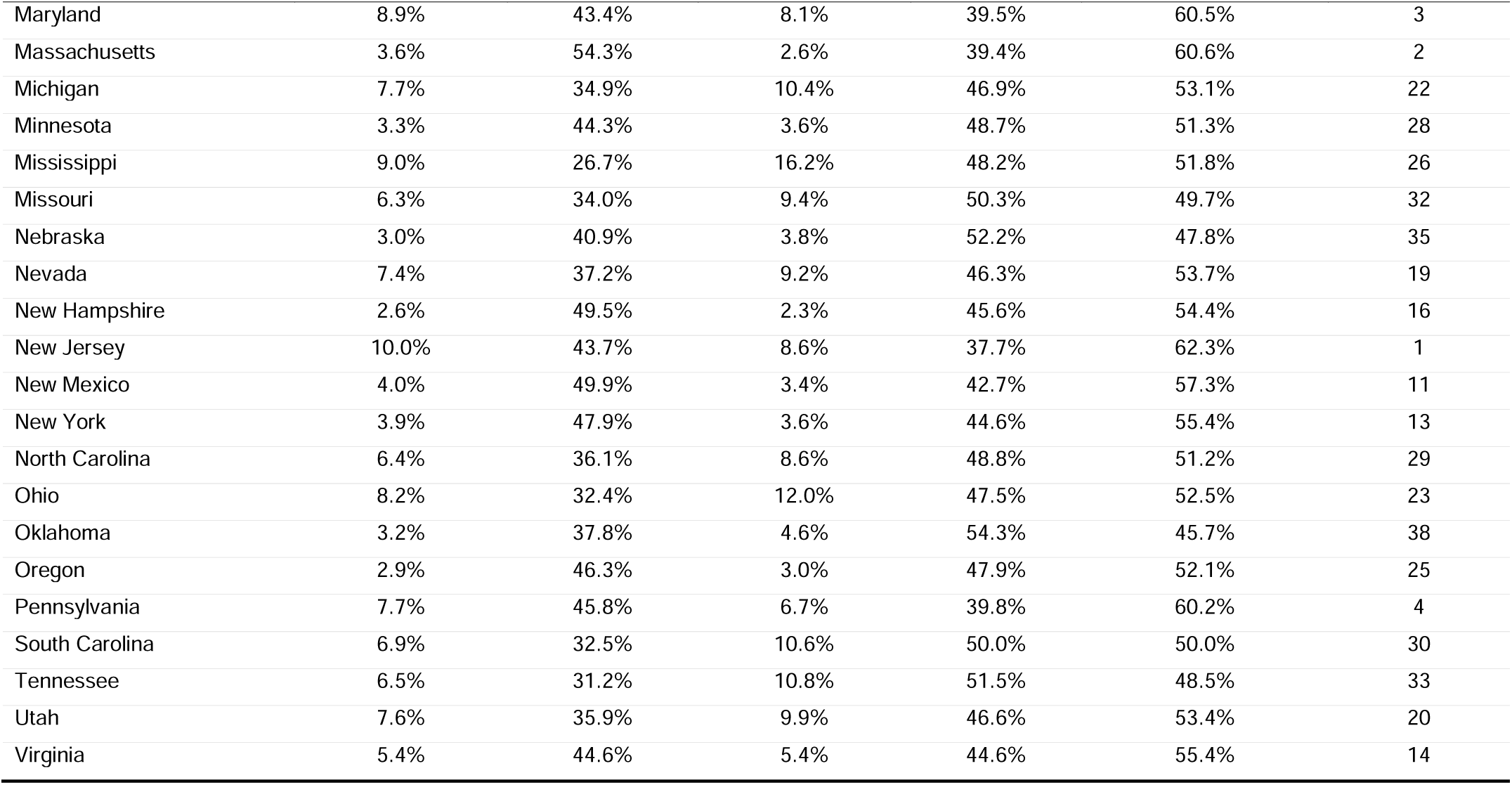
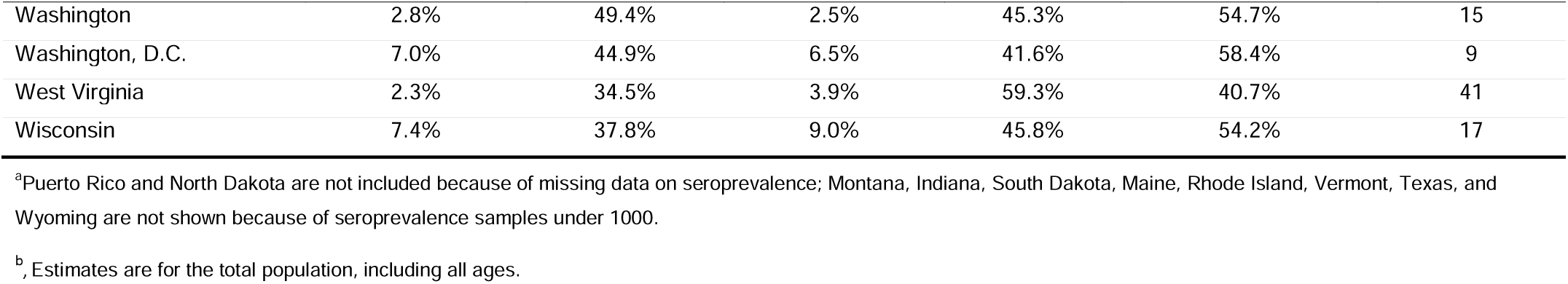
Sensitivity analysis to account for waning immunity in the percentage of the population in each category of presumed immunity due to natural infection, vaccination, both, or neither, by state as of August 26, 2021^a^.

## DISCUSSION

We found that the estimated proportion of the population with presumed immunity to SARS-CoV-2 as of August 26, 2021, varied substantially by state in the US. The states with the highest levels of immunity were those with the highest vaccination rates or those with modest vaccination rates but large proportions immune by infection. While four states may have reached thresholds for population immunity based on less infectious variants (67%), we estimated that none of the states would have reached population immunity for more infectious variants (78% or higher) as of August 26, 2021. As variants that are more infectious may continue to emerge, it is critical that we continue to monitor and intensify vaccination efforts across all states.

The number of states with a high proportion of the population with no presumed immunity to SARS-CoV-2 is high, with proportions reaching >50% in four states and ≥40% in 19 states. The findings with no presumed immunity to SARS-CoV-2 portend continued cases, hospitalizations, and large outbreaks of COVID-19. Further, after accounting for waning immunity, we found that presumed immunity may be even lower, which would lead to an even higher likelihood for sustained transmission than in our original calculations.

With the low levels of presumed immunity in many states, increasing the immunity of the population by vaccinating children will be important. In 2019, 14.6% of the US population was aged <12 years. [26] Achieving population immunity levels >75% for the entire population (all ages) will be difficult without achieving substantial immunity in children.

There were several limitations to our approach. First, we used data from CDC’s Nationwide Commercial Laboratory Seroprevalence Surveys because they provide nucleocapsid seroprevalence estimates for each state. As these surveys use blood samples submitted for reasons unrelated to COVID-19 (e.g., routine clinical visits), they might not be representative of the broader U.S. population. Specifically, people who have blood samples taken during routine medical care or sick visits sought health care and had a blood test and, therefore, may differ in their overall health and disease exposure risk from the general US population.[16] If these individuals are more likely to have other conditions or exposures that also put them at increased risk for COVID-19, this could result in a higher seroprevalence estimate from the survey population being applied to the general US population in our analysis.

Second, although we conducted a sensitivity analysis to explore the potential impact of waning immunity, we were unable to more precisely account for waning immunity after infection or vaccination due to a lack of data on date of infection and limited information about duration of natural immunity. Emerging data suggest that immunity due to infection and vaccination may wane over time and may depend on factors including individual characteristics and the SARS-CoV-2 variant. Although this could result in an overestimate of immunity in our calculations (both primary and sensitivity analyses), our use of recent seroprevalence data should be a reasonably accurate and up-to-date proxy for current immunity in the population. Third, to calculate the percentage of the population with immunity, we assumed that immunity due to infection and vaccination were independent. It is possible that vaccination status might vary by infection status, if individuals who had experienced COVID-19 were more or less likely to seek vaccination than those who had not. We were unable to find representative estimates in the literature describing this relationship. Fourth, this was a cross-sectional analysis, conducted at a single point in time, and therefore could not incorporate changes over time or newer emerging variants. A more comprehensive model would be needed to incorporate these additional factors. Lastly, there are limitations with conducting this analysis at the state level. We were unable to identify relevant seroprevalence estimates by county, and substantial heterogeneity in both disease incidence and vaccine uptake exists for numerous reasons, including population characteristics, population density, and vaccine acceptance. While overall immunity at the larger population levels is critical, the ability to control transmission is also highly dependent on local levels of immunity (e.g., within cities and communities). Geographic clustering of low vaccination rates and/or limited natural immunity will provide opportunities for sustained transmission.[27–29] More sophisticated approaches should monitor changes over time and by different levels of geography to help inform vaccination outreach and guide policy decisions.

In summary, the percentage of the population with presumed immunity varied substantially across the United States as of August 26, 2021. States with the highest calculated levels of presumed immunity had the highest vaccination rates. A small number of states had population immunity over the minimum threshold for less infectious variants (67%), but no states were estimated to have reached population immunity using a threshold for more infectious variants (78% or higher) at the time of this analysis. A third or greater of the population in all states have neither vaccine-derived nor natural immunity to SARS-CoV-2; therefore, it is imperative that vaccination efforts intensify within all states and that uptake occurs across all ages as soon as vaccines are available. Tracking variation among states and counties is important for public health response and ensuring that resources are targeted to communities at the highest risk of amplified SARS-CoV-2 transmission. Without increased population immunity, risk of continued transmission and large outbreaks will exist throughout the US.

## Data Availability

Data available by contacting the corresponding author

## Author contributions

MCDS led writing and data analysis for the article. All other authors contributed to the design of the study, review of analysis, interpretation of results, writing and reviewing and editing the manuscript.

## Acknowledgements

The authors thank Kibri Everett of RTI International for creating the maps used in this article and David Harris and John Forbes of RTI Health Solutions for manuscript review and manuscript editing, respectively.

